# Exploring the Capabilities of ChatGPT in Women’s Health

**DOI:** 10.1101/2024.02.27.23300005

**Authors:** Magdalena Elisabeth Bachmann, Ioana Duta, Emily Mazey, William Cooke, Manu Vatish, Gabriel Davis Jones

## Abstract

**Introduction:** Artificial Intelligence (AI) is redefining healthcare, with Large Language Models (LLMs) like ChatGPT offering novel and powerful capabilities in processing and generating human-like information. These advancements offer potential improvements in Women’s Health, particularly Obstetrics and Gynaecology (O&G), where diagnostic and treatment gaps have long existed. Despite its generalist nature, ChatGPT is increasingly being tested in healthcare, necessitating a critical analysis of its utility, limitations and safety. This study examines ChatGPT’s performance in interpreting and responding to international gold standard benchmark assessments in O&G: the RCOG’s MRCOG Part One and Two examinations. We evaluate ChatGPT’s domain- and knowledge area-specific accuracy, the influence of linguistic complexity on performance and its self-assessment confidence and uncertainty, essential for safe clinical decision-making.

**Methods:** A dataset of MRCOG examination questions from sources beyond the reach of LLMs was developed to mitigate the risk of ChatGPT’s prior exposure. A dual-review process validated the technical and clinical accuracy of the questions, omitting those dependent on previous content, duplicates, or requiring image interpretation. Single Best Answer (SBA) and Extended Matching (EMQ) Questions were converted to JSON format to facilitate ChatGPT’s interpretation, incorporating question types and background information. Interaction with ChatGPT was conducted via OpenAI’s API, structured to ensure consistent, contextually informed responses from ChatGPT. The response from ChatGPT was recorded and compared against the known accurate response. Linguistic complexity was evaluated using unique token counts and Type-Token ratios (vocabulary breadth and diversity) to explore their influence on performance. ChatGPT was instructed to assign confidence scores to its answers (0–100%), reflecting its self-perceived accuracy. Responses were categorized by correctness and statistically analysed through entropy calculation, assessing ChatGPT’s capacity for self-evaluating certainty and knowledge boundaries.

**Findings:** Of 1,824 MRCOG Part One and Two questions, ChatGPT’s accuracy on MRCOG Part One was 72.2% (95% CI 69.2–75.3). For Part Two, it achieved 50.4% accuracy (95% CI 47.2–53.5) with 534 correct out of 989 questions, performing better on SBAs (54.0%, 95% CI 50.0–58.0) than on EMQs (45.0%, 95% CI 40.1–49.9). In domain-specific performance, the highest accuracy was in Biochemistry (79.8%, 95% CI 71.4–88.1) and the lowest in Biophysics (51.4%, 95% CI 35.2–67.5). The best-performing subject in Part Two was Urogynaecology (63.0%, 95% CI 50.1–75.8) and the worst was Management of Labour (35.6%, 95% CI 21.6–49.5). Linguistic complexity analysis showed a marginal increase in unique token count for correct answers in Part One (median 122, IQR 114–134) compared to incorrect (median 120, IQR 112–131, p=0.05). TTR analysis revealed higher medians for correct answers with negligible effect sizes (Part One: 0.66, IQR 0.63–0.68; Part Two: 0.62, IQR 0.57–0.67) and p-values ***<***0.001. Regarding self-assessed confidence, the median confidence for correct answers was 70.0% (IQR 60–90), the same as for incorrect choices identified as correct (p***<***0.001). For correct answers deemed incorrect, the median confidence was 10.0% (IQR 0–10), and for incorrect answers accurately identified, it was 5.0% (IQR 0–10, p***<***0.001). Entropy values were identical for correct and incorrect responses (median 1.46, IQR 0.44–1.77), indicating no discernible distinction in ChatGPT’s prediction certainty.

**Conclusions:** ChatGPT demonstrated commendable accuracy in basic medical queries on the MRCOG Part One, yet its performance was markedly reduced in the clinically demanding Part Two exam. The model’s high self-confidence across correct and incorrect responses necessitates scrutiny for its application in clinical decision-making. These findings suggest that while ChatGPT has potential, its current form requires significant refinement before it can enhance diagnostic efficacy and clinical workflow in women’s health.

## Introduction

Artificial Intelligence (AI) has emerged as a transformative technology in healthcare. At the forefront of this AI revolution are Large Language Models (LLMs), powerful systems designed to mimic human language processing abilities. These LLMs, trained on vast volumes of data encompassing books, articles, websites and other media possess the potential to drive advancements in medicine, a field where precision and safety are paramount. Chat Generative Pre-trained Transformer (ChatGPT) has recently emerged as the prominent LLM. ChatGPT, first released to the public in November 2022 by OpenAI, represents a significant advancement in the field of Natural Language Processing (NLP).[1] This large-scale, multimodal model is adept at understanding and generating human-like text, making it a potentially valuable tool in medicine and healthcare.[2, 3]

Women’s health, specifically Obstetrics and Gynaecology (O&G) is a medical domain poised to derive significant benefit. O&G, a field with a history of significant diagnostic and treatment gaps, could leverage LLMs to bridge these disparities.[4–8] AI could aid in analysing patient histories, imaging, and test results to assist in early and accurate diagnoses. Additionally, AI-driven tools could provide personalized treatment options by processing large datasets to predict the most effective interventions for individual patients. The utilisation of LLMs in O&G not only offers the potential to enhance patient outcomes but also democratise healthcare knowledge, narrowing the existing health inequity gap.

However, the benefits of ChatGPT must be tempered by an acute awareness of its limitations, especially within the complex landscape of healthcare. ChatGPT has been described as a “jack of all trades, master of none”.[9] Nonetheless, it is already being explored by doctors and patients as an adjunct to the traditional medical pathway.[10–14] Ethical concerns regarding this technology are more prevalent than ever, encompassing issues of bias, information governance, patient confidentiality, transparency and accountability.[15, 16] ChatGPT’s propensity to generate content that is convincing yet factually incorrect, often referred to as “hallucinations,” further complicates its potential utility in medical settings. The model’s inability to provide a rationale for erroneous decisions further complicates matters, raising concerns about safety, interpretability, reproducibility and the handling of uncertainty, all of which could have profound implications for patients. While ChatGPT holds immense promise, its application in healthcare requires a careful and thorough evaluation to ensure both its reliability and its limitations are understood.

The O&G specialty training programme in the UK is a structured, continuous educational path that spans seven years. It combines both basic and advanced training stages.[17] Training begins after a doctor has completed their initial medical training, gained foundational competencies over two years of work and achieved full registration with the General Medical Council (GMC).[18] During the programme, trainees are required to pass three key exams (MRCOG Parts 1, 2, and 3) at different stages, which assess their clinical knowledge, reasoning and skills in O&G.[19] These exams are also formal requirements in other international O&G training programmes, with over 100 MRCOG test centres outside the UK.[20, 21]

The MRCOG Part 1 exam is designed to assess trainees’ foundational scientific knowledge. This exam covers four key knowledge domains: cell function, human structure, measurement and manipulation, and understanding illness, encompassing various subjects including physiology, anatomy, biophysics, and clinical management.[22] The MRCOG Part 2 exam advances the assessment to a more practical level, testing the application of the knowledge acquired, i.e. clinical reasoning.[22] It comprises a mixture of single best answer (SBA) and extended matching questions (EMQ). These evaluate the trainee’s theoretical understanding as well as their ability to apply this knowledge in practical scenarios. The combination of these question types ensures a comprehensive assessment of the trainee’s capabilities in O&G, preparing them for advanced practice in the field. The MRCOG exams hold significant international recognition and are widely regarded as a gold standard qualification in O&G. Achieving the MRCOG qualification after a medical degree is regarded as a benchmark of medical competence.

The nature of questions found in the MRCOG examinations, specifically SBAs and EMQs, provide a pertinent framework for gauging the capabilities of LLMs such as ChatGPT. These formats are particularly challenging because they often present multiple answers that could all be considered correct. Clinicians must draw upon not only their knowledge, but also their clinical reasoning and experience to discern the most appropriate answer from among various plausible options. Thus, when ChatGPT is tasked with identifying the single best answer, it undergoes a rigorous test of its clinical reasoning abilities. This goes beyond simple recollection of information, requiring instead the application of knowledge to a defined clinical context, as per the standards established by the RCOG and accepted clinical practice.

The objectives of this study were threefold: Firstly, to assess the efficacy of ChatGPT in interpreting and responding to questions from the MRCOG Part 1 and Part 2 examinations, thus evaluating its domain-specific accuracy in a standardised medical knowledge and reasoning context. Secondly, to determine whether the complexity of the questions influences ChatGPT’s performance accuracy, thereby enabling an analysis of its clinical knowledge and reasoning capabilities independent of linguistic difficulty. Thirdly, to investigate ChatGPT’s self-assessment of confidence in its responses, providing insight into the reliability and safety of AI in clinical decision-making processes. This self-evaluation aspect is particularly crucial, as it could reflect the model’s ability to estimate its certainty and, by extension, its utility in real-world medical applications where the cost of error is potentially high.

## Methods

### Data Acquisition and Processing

We extracted single best answer (SBA) and extended matching questions (EMQ) questions for the MRCOG Part One and Part Two examinations from online sources regarded as unavailable to LLMs trained on publicly available data. This was done to reduce the possibility of evaluating ChatGPT on examination questions it had already observed and memorised. Sources included the Royal College of Obstetrics and Gynaecology and publishers making their content only available to users with the appropriate license.[23] Data extraction was permitted under the exception of Section 29 of the UK Copyright, Designs and Patents Act 1988 which allows researchers to make copies of copyright works for non-commercial research. Only questions from 2015 onwards were used to avoid including questions that may have been used in the training of ChatGPT or no longer adhered to current clinical practice guidelines. Prior to inclusion in the study database, each question and corresponding answers underwent validation. This involved a dual-review system where a data scientist ensured the technical accuracy of the conversion to the study format and the clinical team confirmed the medical accuracy and relevance. Questions that relied on information from previous questions were updated to include that information, while those that were duplicated or included images for interpretation were omitted.

Questions were converted into a format for simplified interpretation by ChatGPT. We chose the JavaScript Open Notation (JSON) format for its flexibility and widespread use in data interchange.[24] JSON’s hierarchical structure allows for the representation of complex question and answer formats, facilitating the efficient parsing of data by ChatGPT. To ensure tabular data retained its context and was interpretable, we developed a conversion protocol that preserved the relational structure of tables, converting them into nested JSON objects that ChatGPT could systematically evaluate. The background information for the examination (including the type and nature of question being asked, e.g. SBA or EMQ) was incorporated into the instruction given to ChatGPT. The knowledge area and domain of understanding assigned by the publisher for each question was recorded with the question for sub-analysis (e.g. anatomy, biophysics, urogynaecology). Where the subject was not provided by the source, these were assigned by the clinical team.

Interfacing with the OpenAI application programming interface (API) was accomplished using a Python script.[25] We ensured that each query to the API was structured to mirror the interactive nature of the ChatGPT interface, including the provision of context where necessary and the structured format of the JSON-encoded data. Each prompt for ChatGPT was developed in accordance with prompt engineering guidelines.[26, 27] Parameters such as temperature, which controls the randomness of the response, were set to zero to favour deterministic outputs, providing consistency across multiple requests. The complete prompt was then provided to ChatGPT and the responses recorded. ChatGPT was presented with each prompt individually to avoid contamination of responses, ensuring that each response was generated based on the input provided without influence from neighbouring questions. ChatGPT was not subsequently informed of the correct answer. The response was then compared against the correct answer for each question.

### Linguistic complexity analysis

We then investigated the role of linguistic complexity in model performance. Each question was tokenised and metrics including unique token count and type-token ratio (TTR) were computed.[28] The unique token count represented the total number of distinct words used (the breadth of vocabulary) while the TTR provided a measure of lexical diversity (the diversity of that vocabulary relative to the total number of words used). These were selected as metrics of linguistic complexity because they offer insights into the variety and richness of the language used within the questions. These metrics are indicative of the complexity ChatGPT must navigate to understand and respond to a question, hypothesizing that a higher linguistic complexity might affect ChatGPT’s performance.

### Self-assessed confidence and uncertainty

Finally, we aimed to determine the extent to which ChatGPT could self-assess the confidence and uncertainty of its responses. We conducted a series of experiments wherein ChatGPT was instructed to assign a probabilistic confidence score, ranging from 0—1 (0—100%), to each answer option within a question. The decision to assess confidence using a probability score is grounded in the probabilistic nature of ChatGPT’s language model. These confidence scores were then utilised as an indicator of the model’s self-perceived accuracy when the correct answer was identified. The responses deemed incorrect by ChatGPT were bifurcated into two categories: those incorrectly classified as erroneous and those accurately classified as such. A higher confidence score was interpreted as indicative of greater certainty in the response.

Entropy was calculated for the distribution of confidence scores to quantitatively measure the model’s uncertainty. Entropy was calculated using the Shannon entropy formula, a fundamental concept in information theory that measures the unpredictability or randomness of information content.[29] In this context, it quantifies the degree of uncertainty in ChatGPT’s predictions. Entropy values inversely correlate with uncertainty; thus, lower entropy signifies greater confidence in the responses, and higher entropy indicates greater uncertainty. This analysis provided a statistical layer to the confidence scores, enriching our understanding of the model’s performance. A statistically significant difference in confidence or uncertainty levels across different categories would imply an intrinsic capability of ChatGPT to discern the boundaries of its knowledge within specific domains.

### Statistical Analysis

Categorical variables are expressed as frequencies and percentages. Continuous variables not normally distributed are described using medians and interquartile ranges (IQR). The accuracy metric was defined as the ratio of correct predictions to total predictions made by ChatGPT, where a correct prediction is denoted as a congruence between ChatGPT’s prediction and the true value (e.g., both ChatGPT prediction and correct answer are ’A’). Accuracy and probability values are reported as percentages. Differences between categorical variables were assessed with the Chi-square test. Continuous variables were evaluated using the Mann-Whitney U test, considering a p-value of less than 0.05 as statistically significant. The analysis utilised the GPT-4 model (”gpt-4”, accessed 9th November, 2023) to assess responses to queries. All statistical computations were conducted with Python (version 3.9.17), employing libraries including Pandas (version 1.5.3), NumPy (version 1.23.5), Matplotlib (version 3.7.1), OpenAI (version 0.28.1), and TikToken (version 0.5.1).

## Results

1,824 MRCOG Part One and Part Two questions from eight sources were extracted and converted into a format readable for ChatGPT. 835 MRCOG Part One single best answer (SBA) questions and 989 MRCOG Part Two questions (589 SBAs and 400 extended matching questions [EMQ]) were identified. The range of answer choices for SBA questions was between A–E (5 options) while the range for EMQs was between 5–18 choices (A–R). 56 questions (3.1%) contained additional tabular data which were converted to JSON format. 4 questions with associated images were omitted. Questions were identified for each of the areas of knowledge prescribed by the RCOG (14 for Part One and 15 for Part Two examinations). The median number of questions in each knowledge area for Parts One and Two were 58 (IQR 32–85) and 45 (IQR 32–64). See Supplementary Tables 1 & 2 for the distribution of knowledge areas.

### ChatGPT Performance Accuracy

ChatGPT achieved an overall accuracy of 72.2% (95% CI 69.2–75.3, 603/835 correct) on Part One and 50.4% (95% CI 47.2–53.5, 534/989 correct) on Part Two of the MRCOG examinations. Across the four domains of understanding for the MRCOG Part One examination (Table 1, Figure 1), there was a significant difference in the accuracy of ChatGPT (p=0.02, *χ*^2^= 9.85). ChatGPT performed best in the “Illness” domain with an accuracy of 80.0% (95% CI 73.3–85.7) and worst in the “Measurement and Manipulation” domain with an accuracy of 65.7% (95% CI 58.8–72.7). We then evaluated the accuracy of ChatGPT in the subjects constituting these domains (Table 2, Figure 1). There was no significant difference between each subject within any domain (Domain-specific p-values: Cell Function, p=0.08; Human Structure, p=0.07; Illness, p=0.49; Measurement and Manipulation, p=0.11, Table 2). For each domain, ChatGPT demonstrated the highest accuracy in Biochemistry (79.8% [95% CI 71.4–88.1], Cell Function), Embryology (80.4% [95% CI 70.0–90.8], Human Structure), Clinical Management (83.3% [95% CI 68.4–98.2], Understanding Illness) and Pharmacology (75.4% [95% CI 64.3–86.6], Measurement and Manipulation). The subjects ChatGPT performed worst in within each domain were Physiology (65.3% [95% CI 56.1–74.6], Illness), Anatomy (63.2% [95% CI 54.0–72.4], Human Structure), Immunology (70.0% [95% CI 53.6–86.4], Illness) and Biophysics (51.4% [95% CI 35.2–67.5], Measurement and Manipulation).

**Fig. 1:**
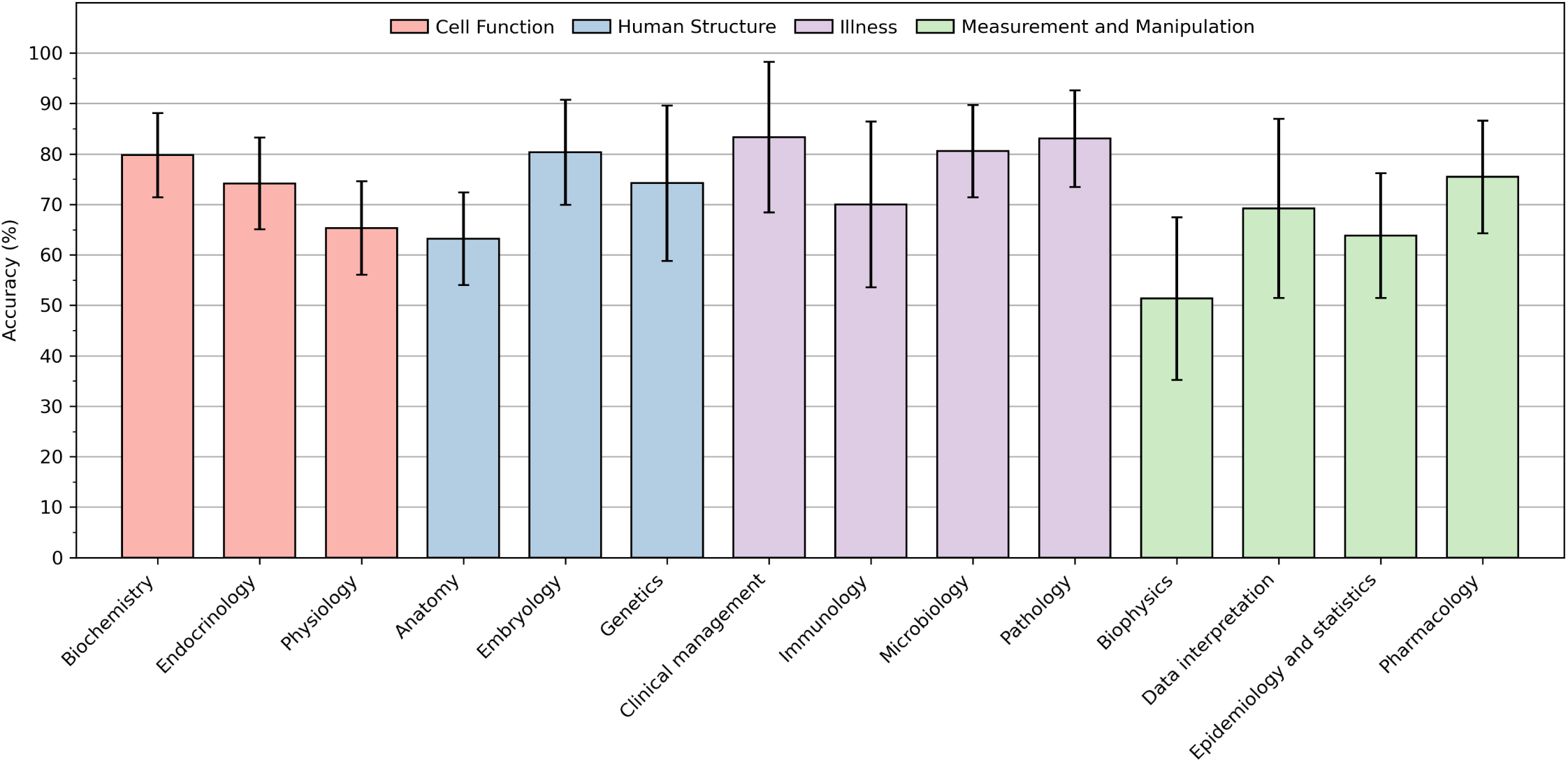
Performance of ChatGPT on the MRCOG Part One examination. Significant variance in performance across the four domains was noted (p=0.02; *χ*^2^= 9.85). The highest accuracy was observed in the domain of “Illness” at 79.5% (95% Confidence Interval [CI]: 73.3–85.7), whereas the lowest was in “Measurement and Manipulation” at 65.7% (95% CI: 58.8–72.7). Analysis of ChatGPT’s accuracy within individual subjects corresponding to these domains revealed no substantial differences (Domain-specific p-values: Cell Function, p=0.08; Human Structure, p=0.07; Illness, p=0.49; Measurement and Manipulation, p=0.11). Within each domain, subjects with the highest accuracy were Biochemistry (79.8% [95% CI: 71.4–88.1], Cell Function), Embryology (80.4% [95% CI: 70.0–90.8], Human Structure), Clinical Management (83.3% [95% CI: 68.4–98.2], Illness), and Pharmacology (75.4% [95% CI: 64.3–86.6], Measurement and Manipulation). Subjects with the lowest accuracy were Physiology (65.3% [95% CI: 56.1–74.6], Illness), Anatomy (63.2% [95% CI: 54.0–72.4], Human Structure), Immunology (70.0% [95% CI: 53.6–86.4], Illness), and Biophysics (51.4% [95% CI: 35.2–67.5], Measurement and Manipulation).

**Table 1:** ChatGPT Performance Accuracy across the Four Domains of the MRCOG Part One examination. The overall accuracy was 72.2% (95% CI 69.2–75.3). There was a significant difference in the accuracy of ChatGPT across the four domains (p=0.02, *χ*^2^ statistic = 9.85). ChatGPT performed best in the “Illness” domain with an accuracy of 80.0% (95% CI 73.3–85.7) and worst in the “Measurement and Manipulation” domain with an accuracy of 65.7% (95% CI 58.8–72.7). Values in brackets denote the percentage proportion (%).

| Domain | Correct | Incorrect | Total |
| --- | --- | --- | --- |
| Cell Function | 203 (72.8%) | 76 (27.2%) | 279 |
| Human Structure | 135 (69.9%) | 58 (30.1%) | 193 |
| Illness | 148 (80.0%) | 37 (20.0%) | 185 |
| Measurement and Manipulation | 117 (65.7%) | 61 (34.3%) | 178 |
| **Total** | **603 (72.2%)** | **232 (27.8%)** | **835** |

**Table 2:** ChatGPT Performance Accuracy in each Subject comprising the MRCOG Part One domains. There was a significant difference in the accuracy of ChatGPT across the four domains (p=0.02, *χ*^2^ = 9.85), however the performance of each subject within any domain was not significantly different. Values in brackets denote the percentage proportion (%).

| Domain | Knowledge area | Correct | Incorrect | Total | p-value |
| --- | --- | --- | --- | --- | --- |
| Cell Function | Biochemistry | 45 (80.4%) | 11 (19.6%) | 56 | 0.08 |
|  | Endocrinology | 67 (63.2%) | 39 (36.8%) | 106 |  |
|  | Physiology | 23 (74.2%) | 8 (25.8%) | 31 |  |
| Human Structure | Anatomy | 71 (79.8%) | 18 (20.2%) | 89 | 0.07 |
|  | Embryology | 66 (74.2%) | 23 (25.8%) | 89 |  |
|  | Genetics | 66 (65.3%) | 35 (34.7%) | 101 |  |
| Illness | Clinical management | 37 (63.8%) | 21 (36.2%) | 58 | 0.49 |
|  | Immunology | 19 (51.4%) | 18 (48.6%) | 37 |  |
|  | Microbiology | 43 (75.4%) | 14 (24.6%) | 57 |  |
|  | Pathology | 18 (69.2%) | 8 (30.8%) | 26 |  |
| Measurement & Manipulation | Biophysics | 21 (70.0%) | 9 (30.0%) | 30 | 0.11 |
|  | Data interpretation | 49 (83.1%) | 10 (16.9%) | 59 |  |
|  | Epidemiology and statistics | 58 (80.6%) | 14 (19.4%) | 72 |  |

For Part Two, the RCOG does not assign subjects to discrete domains, as subjects and questions can span multiple domains. Therefore ChatGPT’s performance was assessed by subject only. The accuracy across subjects did not vary significantly (p=0.10, *χ*^2^= 21.05, Table 3, Figure 2). The best performing knowledge area was Urogynaecology & Pelvic Floor Problems (accuracy 63.0% [95% CI 50.1–75.8]) while the worst performing area was Management of Labour (accuracy 35.6% [95% CI 21.6–49.5]. ChatGPT performed better at SBA questions (54.0% accurate [95% CI 50.0-58.0]) than EMQ questions (45.0% accurate [95% CI 40.1–49.9], p=0.01, *χ*^2^=7.35, Table 4).

**Fig. 2:**
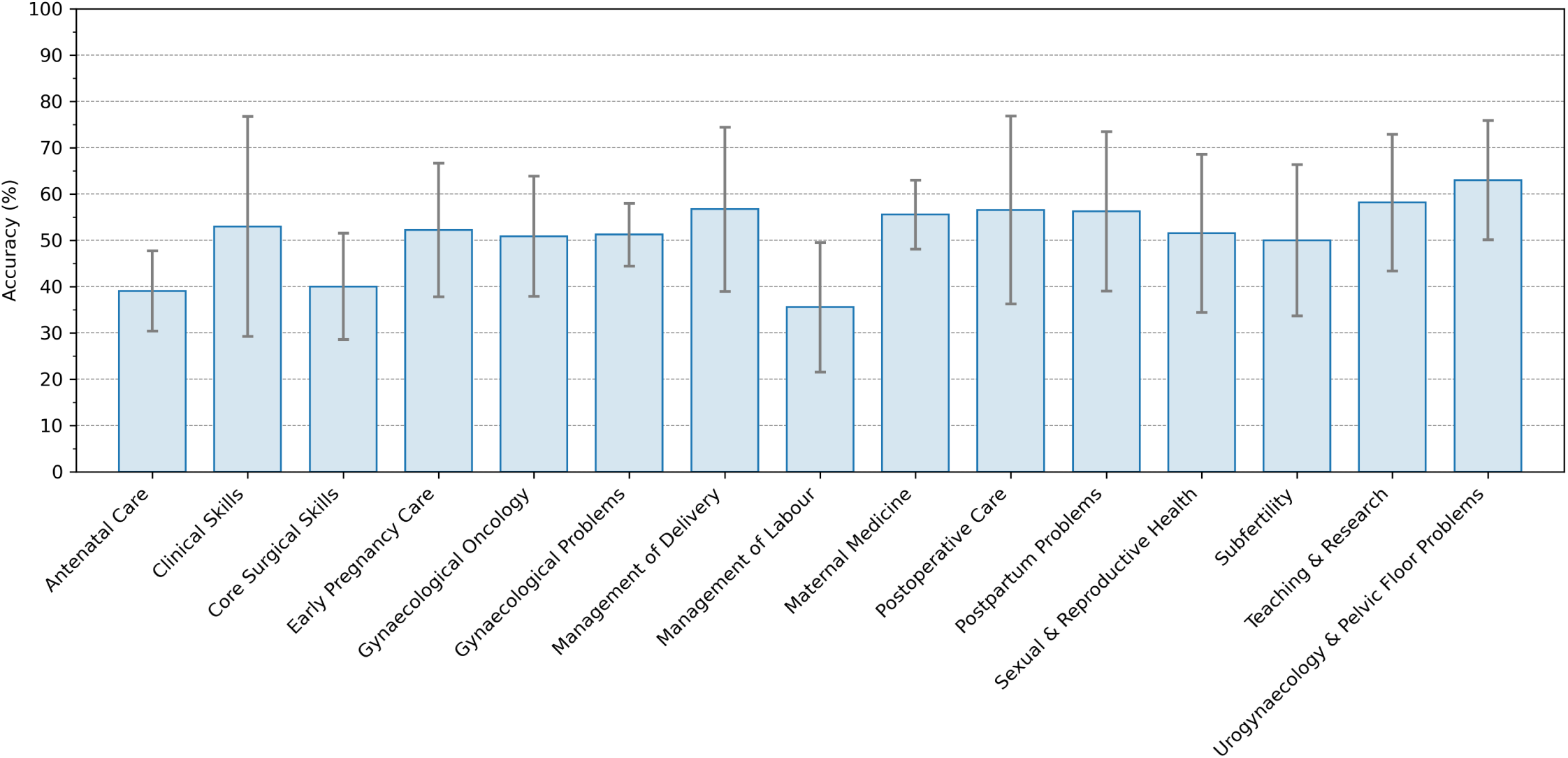
Performance of ChatGPT on the MRCOG Part Two examination. The distribution of accuracies across various knowledge areas showed no significant variation (p=0.10, *χ*^2^= 21.05). Urogynaecology & Pelvic Floor Problems emerged as the area with the highest accuracy at 63.0% (95% Confidence Interval [CI]: 50.1–75.8), contrasting with Management of Labour which had the lowest at 35.6% (95% CI: 21.6–49.5).

**Table 3:** ChatGPT Performance Accuracy in the MRCOG Part Two. Part Two comprises single best answer (SBA) and extended matching questions (EMQ) from 15 knowledge areas (subjects). Accuracy across the knowledge areas did not vary significantly (p=0.10, *χ*^2^= 21.05). Values in brackets denote the percentage proportion (%).

| Knowledge area | Correct | Incorrect | Total |
| --- | --- | --- | --- |
| Antenatal Care | 48 (39.0%) | 75 (61.0%) | 123 |
| Clinical Skills | 9 (52.9%) | 8 (47.1%) | 17 |
| Core Surgical Skills | 28 (40.0%) | 42 (60.0%) | 70 |
| Early Pregnancy Care | 24 (52.2%) | 22 (47.8%) | 46 |
| Gynaecological Oncology | 29 (50.9%) | 28 (49.1%) | 57 |
| Gynaecological Problems | 107 (51.2%) | 102 (48.8%) | 209 |
| Management of Delivery | 17 (56.7%) | 13 (43.3%) | 30 |
| Management of Labour | 16 (35.6%) | 29 (64.4%) | 45 |
| Maternal Medicine | 95 (55.6%) | 76 (44.4%) | 171 |
| Postoperative Care | 13 (56.5%) | 10 (43.5%) | 23 |
| Postpartum Problems | 18 (56.2%) | 14 (43.8%) | 32 |
| Sexual & Reproductive Health | 17 (51.5%) | 16 (48.5%) | 33 |
| Subfertility | 18 (50.0%) | 18 (50.0%) | 36 |
| Teaching & Research | 25 (58.1%) | 18 (41.9%) | 43 |
| Urogynaecology & Pelvic Floor Problems | 34 (63.0%) | 20 (37.0%) | 54 |
| **Total** | **498** | **491** | **989** |

**Table 4:** Comparing ChatGPT’s Performance Accuracy between SBAs and EMQs. ChatGPT performed better in single best answer (SBA) questions than extended matching questions (EMQ), *p* = 0.01, *χ*^2^ = 7.35. Values in brackets denote the percentage proportion (%).

| Question Type | Correct | Incorrect | Total |
| --- | --- | --- | --- |
| Single best answer (SBA) | 318 (54.0%) | 271 (46.0%) | 589 |
| Extended matching questions (EMQ) | 180 (45.0%) | 220 (55.0%) | 400 |
| **Total** | **498** | **491** | **989** |

### Influence of Linguistic Complexity on ChatGPT Performance

We next evaluated whether the linguistic complexity of the questions given to ChatGPT could influence its performance. Each question was tokenised and the unique token count and type-token ratio (TTR) were calculated (Table 5). For the MRCOG

**Table 5:**
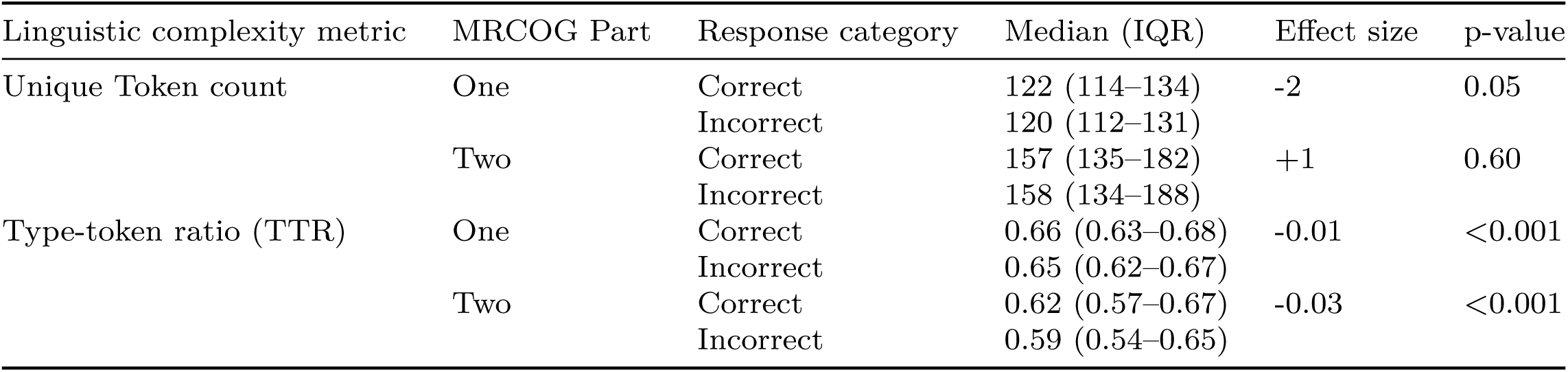
Evaluation of Linguistic Complexity as a Factor in ChatGPT Performance. Values for unique token count have been rounded to whole numbers. Values for TTR have been rounded to 2 significant figures. Effect size describes the average difference between correct and incorrect responses for the linguistic complexity metrics. There was a significant difference observed for the unique token count metric for the MRCOG Part One whereby incorrect responses had on average two unique tokens less than correct responses. A significant difference was observed for the type-token ratio, whereby incorrect responses had an average TTR of -0.01 less than correct responses for part one (p*<*0.001) and -0.03 for part two. These results suggest that, while there was a significant difference, the effect size (actual difference in these values) was not of meaningful importance in ChatGPT’s performance accuracy.

Part One, the median unique token count was marginally higher for correct responses (122 [IQR 114–134]) compared to incorrect responses (120 [IQR 112–131]), with a small effect size of –2 and a p-value of 0.05, indicating a statistically significant but minor difference. In Part Two, no significant difference was found in the unique token count between correct and incorrect responses (p=0.60). A statistically significant difference was observed for TTR. In Part One, correct responses had a slightly higher median TTR (0.66 [IQR 0.63–0.68]) compared with incorrect responses (0.65 [IQR 0.62–0.67]), with a negligible effect size of -0.01 (p*<*0.001). Similarly, for Part Two, correct responses had a median TTR of 0.62 (IQR 0.57–0.67), which was marginally higher than the 0.59 (IQR 0.54–0.65) of incorrect responses, with an effect size of -0.03 (p*<*0.001). These findings suggest that the linguistic complexity, as measured by unique token count and TTR, has a statistically significant association with the accuracy of responses. However, the effect sizes indicate that the actual difference in linguistic complexity between correct and incorrect responses is not substantial enough to meaningfully influence ChatGPT’s performance.

### Confidence and Uncertainty in ChatGPT Responses

In the evaluation of ChatGPT’s self-assessment of confidence, it was observed that for 192 questions, representing 10.5% of the total, ChatGPT allocated an identical probability score to each answer option. These instances were deemed to lack discriminatory power and were excluded from analysis. Of the remaining probabilities, 1,072 were associated with correct answers accurately identified by ChatGPT. Conversely, 567 probabilities pertained to answers incorrectly identified as correct, another 567 were allocated to correct answers erroneously identified as incorrect, and 3,100 probabilities corresponded to answers correctly identified as incorrect (Figure 3). The high value of 3,100 probabilities in this latter category is explained by their being multiple incorrect answers per question. The median confidence level for both correctly identified correct answers and incorrectly identified correct answers was 70.0% (Interquartile Range [IQR]: 60–90, p*<*0.001). For correct answers misclassified as incorrect, the median confidence was 10.0% (IQR: 0–10), whereas for incorrect answers rightly identified as such, the median confidence was 5.0% (IQR: 0–10, p*<*0.001). Despite statistical significance, the practical difference in confidence levels between these groups was minimal.

**Fig. 3:**
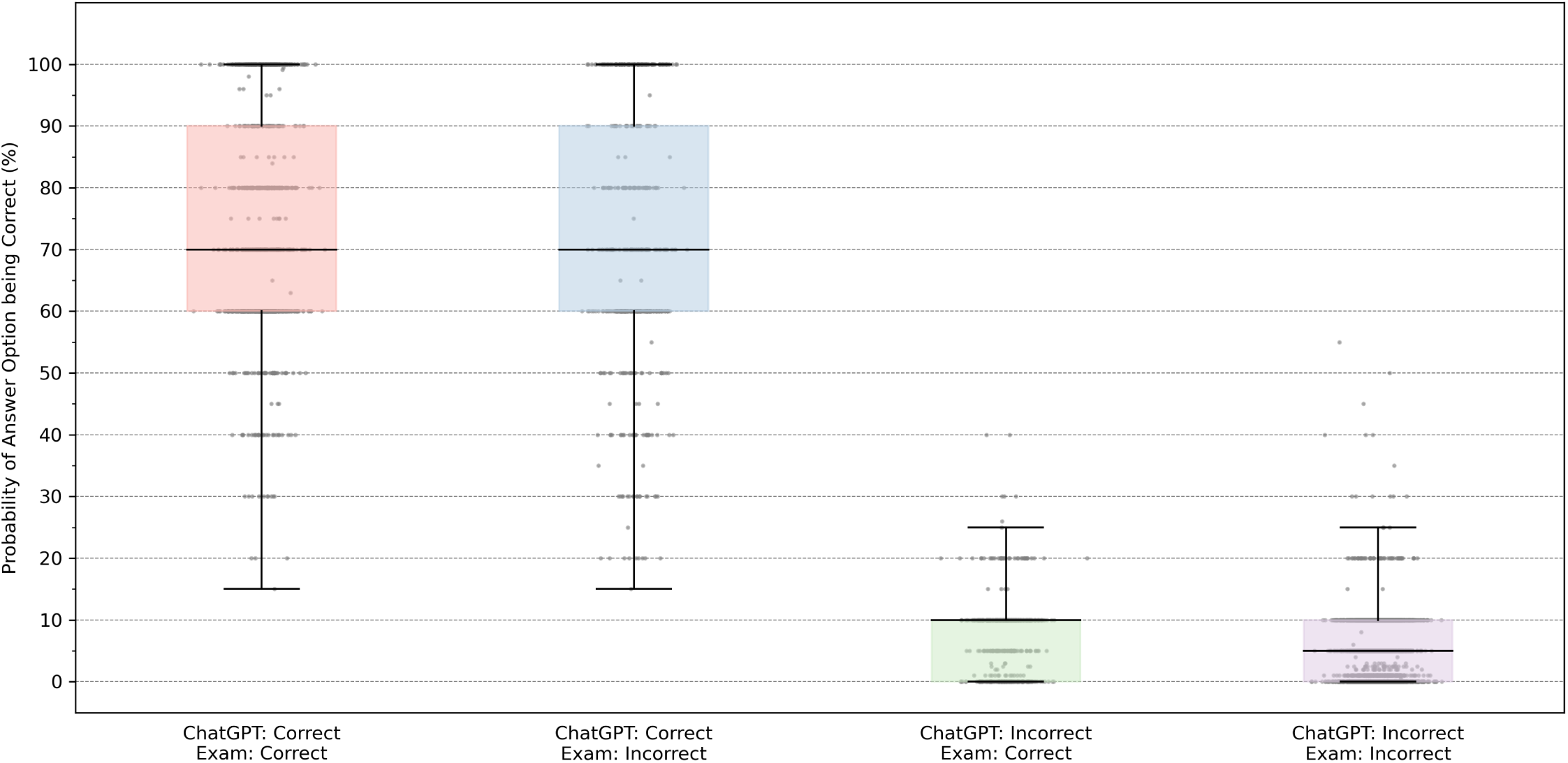
Box-and-Whisker Plot Depicting the Distribution of Confidence Scores Attributed by ChatGPT to Different Categories of Responses. The y-axis represents the probability scores (expressed as a percentage) that ChatGPT assigned to its answers, indicative of self-assessed confidence. The categories on the x-axis represent four scenarios: ChatGPT correctly identifying a correct answer (red), ChatGPT incorrectly identifying a correct answer (blue), ChatGPT incorrectly identifying an incorrect answer as correct (green), and ChatGPT correctly identifying an incorrect answer (purple). The central line in each box denotes the median confidence score, while the bounds of the boxes represent the interquartile range (IQR). Outliers are depicted as individual points. The median confidence level for correctly identified correct answers and incorrectly identified correct answers was both at 70.0%, while for correct answers misclassified as incorrect, the median was significantly lower at 10.0%. Incorrect answers accurately identified as such had a median confidence of 5.0%. Despite the presence of statistical significance, the minimal practical variance in confidence scores suggests a limitation in ChatGPT’s ability to self-evaluate the certainty of its responses accurately.

The median entropy for ChatGPT’s correct responses (where ChatGPT’s answer matched the correct exam answer) was 1.46 (IQR 0.44–1.77) and similarly, the median entropy for its incorrect responses (where ChatGPT’s answer did not match the correct exam answer) was 1.46 (IQR: 0.67–1.77, p*<*0.001, Figure 4). The identical median values suggest that ChatGPT’s distribution of probabilities does not discernibly distinguish between its correct and incorrect responses.

**Fig. 4:**
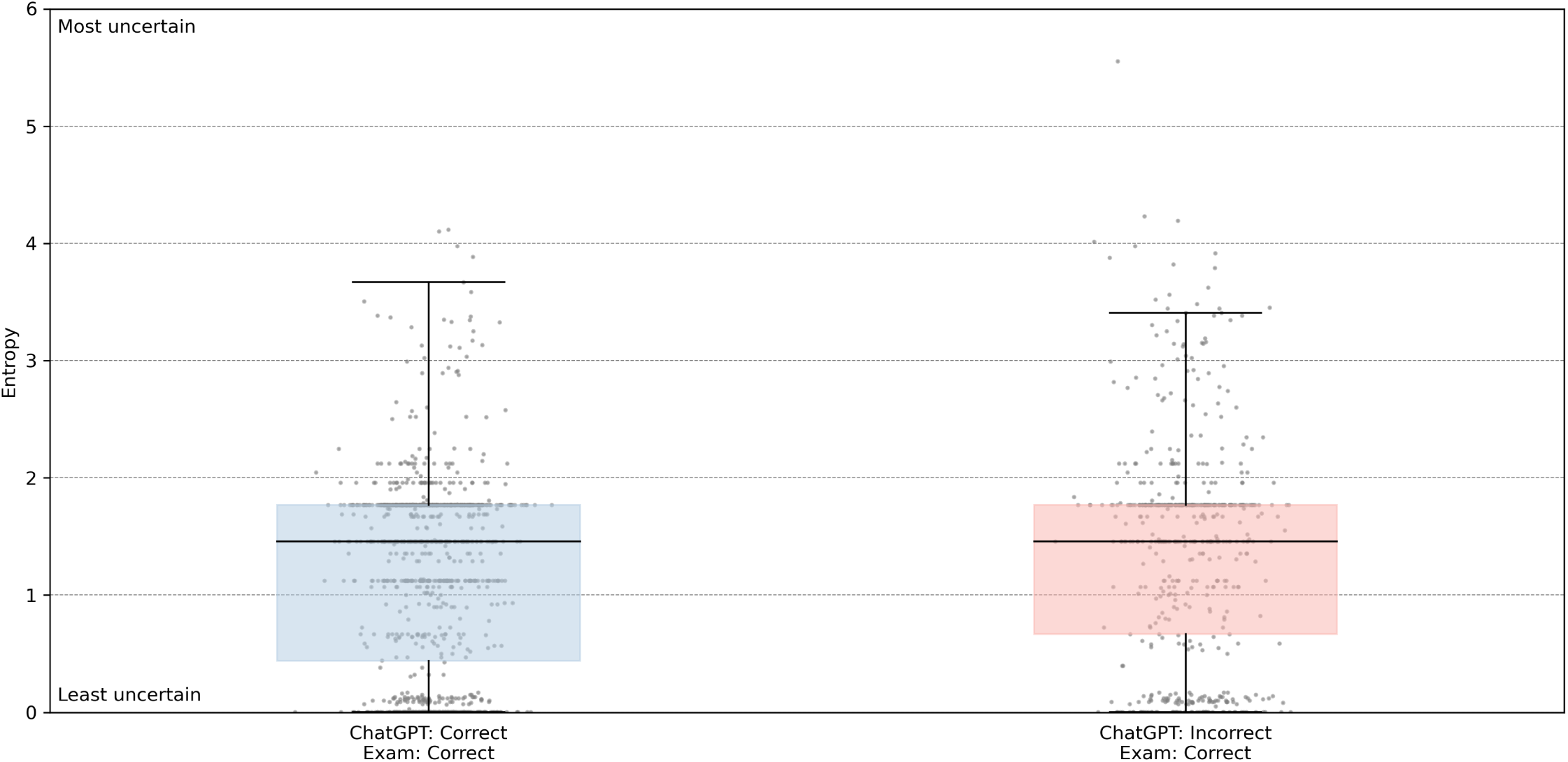
Comparative Entropy Distribution for Correct and Incorrect Responses by ChatGPT. This box-and-whisker plot displays the entropy values for ChatGPT responses, stratified by the model’s accuracy and the actual correctness of the exam answers. The y-axis represents entropy, a measure of uncertainty, with higher values indicating greater uncertainty. The blue box represents responses where ChatGPT’s answers were correct for questions with correct exam answers, showing a median entropy of 1.46 (IQR: 0.44–1.77). The red box denotes responses where ChatGPT’s answers were incorrect for questions with correct exam answers, with an identical median entropy of 1.46 (IQR: 0.67–1.77, p*<*0.001). The consistent median entropy across both categories indicates that ChatGPT’s confidence does not significantly vary between its correct and incorrect responses, despite the statistical significance, calling into question the model’s self-assessment accuracy.

## Discussion

This study represents a novel and in-depth evaluation of the potential for LLMs as tools in women’s health, specifically O&G. Leveraging a substantial dataset of questions from the Royal College of Obstetricians and Gynaecologists’ MRCOG Part One and Part Two examinations, we have detailed a comprehensive analysis of ChatGPT’s capabilities in understanding and applying medical knowledge and reasoning to an internationally-recognised standard of excellence. ChatGPT exhibited a notable level of proficiency in the MRCOG Part One examination, displaying an ability to evaluate medical content based on the current MRCOG syllabus.[17] The syllabus covers basic and applied science knowledge necessary for qualified medical professionals before they begin specialty training in O&G. In contrast, the Part Two examination, which tests candidates with several years of training in O&G on the application of their knowledge (i.e. clinical reasoning) to representative clinical scenarios, ChatGPT’s performance was poorer. While ChatGPT outperformed random chance, its responses were, on average, as frequently incorrect as they were correct. This demonstrates several significant limitations not only in its domain knowledge but also the understanding and application of complex clinical knowledge and reasoning. Given this discrepancy in performance between Part One and Part Two, it would be premature to suggest ChatGPT possesses a comparable or useful level of understanding within women’s health.

This conclusion is reinforced when considering ChatGPT’s overall self-reported confidence and certainty in its answers. It displayed a high degree of confidence in incorrect responses, performing poorly when presented with the correct answer as an option, as evidenced in SBA and EMQ formats. Although statistical significance was observed, the practical implications of this finding remain equivocal, necessitating further investigation to ascertain whether ChatGPT possesses an inherent ability to gauge the veracity of its generated answers with any degree of reliability. This indicates that ChatGPT does not have a reliable mechanism for self-evaluating its confidence or certainty, as evidenced by similar scores for both correct and incorrect responses. This misalignment between confidence and correctness raises concerns regarding the reliability of ChatGPT in clinical decision-making or patient information-giving scenarios. Our combined evaluation of not only performance accuracy but of ChatGPT’s self-reported confidence suggest this is not currently a safe tool for use by either clinicians or patients.

There is growing concern globally surrounding AI safety; our findings support this.[30] While LLMs such as ChatGPT undoubtedly possess substantial potential in several domains it has demonstrated significant limitations in medicine and healthcare.[31–34] Impressive performance in one task does not necessarily translate to equivocal performance in others. Users of this technology, both medical practitioners and patients alike, need be aware. As these AI models continue to develop, we hope to see an improvement in women’s health. Women’s health is a field with a significant diagnostic and treatment gap.[5, 6, 8, 35] Caution must be taken that, through these technologies, it does not widen. Safety in the context of women’s health must be a priority. Work is currently underway to develop and evaluate LLMs trained instead on region-specific clinical best practice guidelines. We are also developing a platform for safely testing LLMs based on local and international clinical consensus. Through this work, we hope to see the development of reliable, robust and safe AI models that can be of utility.

There are several important strengths to this study. We evaluated ChatGPT with data unlikely to have been used in its training. This enabled a more direct and robust interrogation of its aptitude in clinical knowledge and reasoning without the associated bias of testing the AI model on previously learned questions and answers. In essence, we have avoided testing a system on an examination it has already memorised, forcing it instead to use its current domain-specific knowledge and reasoning. We have also evaluated different levels of expected clinical aptitude by examining ChatGPT on Parts One and Two of the MRCOG. Our evaluation encompassed not only the accuracy of responses but the model’s linguistic processing capabilities and its self-assessment of confidence and certainty. We have demonstrated that the poor performance of ChatGPT is not attributable to linguistic complexity. Likewise, we have shown that ChatGPT is equally as confident when it is wrong as when it is correct. Currently, ChatGPT will answer most questions, with relative disregard for safety or accuracy beyond a generic disclaimer. This study was limited in that it did not compare ChatGPT’s performance directly against the performance of candidates undertaking the same examinations – these data are not provided by the RCOG. We posit, however, that LLMs with the potential demonstrated by ChatGPT need to demonstrate at least near-perfect performance. Especially if they are to be made as publicly available as ChatGPT.

In light of our findings, we suggest that for LLMs to be viable in medical practice, they must first unequivocally demonstrate domain competence in both knowledge and reasoning. Such competence entails not only matching (or surpassing) human experts in clinical knowledge and reasoning tasks, which in itself is insufficient to capture the complexities of clinical medicine, but also possessing an acute awareness of the AI’s own boundaries of knowledge and the associated risks when these boundaries are approached or breached.

## Conclusions

In conclusion, the current state of publicly-available LLMs, as exemplified by ChatGPT, while impressive, does not meet the essential criteria for utility in clinical settings or by patients seeking medical information in the domain of Women’s Health. Far from being a ready-to-implement tool, its deficiencies in accuracy and awareness regarding its own limitations, combined with the potential for misinformation and patient risk, render it unsuitable for use in women’s health in its present form.

## Data Availability

The data used in this study are proprietary and subject to copyright and therefore unavailable.

## Notes

### Competing Interest Statement

The authors have declared no competing interest.

### Funding Statement

This study did not receive any funding.

