## Supplementary Information for "Exploring the Capabilities of ChatGPT in Women’s Health"

| MRCOG Part One Knowledge area | Questions sourced |
|---|---|
| Anatomy | 106 |
| Biochemistry | 89 |
| Biophysics | 37 |
| Clinical management | 24 |
| Data interpretation | 26 |
| Embryology | 56 |
| Endocrinology | 89 |
| Epidemiology and statistics | 58 |
| Genetics | 31 |
| Immunology | 30 |
| Microbiology | 72 |
| Pathology | 59 |
| Pharmacology | 57 |
| Physiology | 101 |

Supplementary Figure 1: Distribution of Number of Assessed MRCOG Part One Questions by Knowledge Area.

| MRCOG Part Two Knowledge area | Questions sourced |
|---|---|
| Antenatal Care | 123 |
| Clinical Skills | 17 |
| Core Surgical Skills | 70 |
| Early Pregnancy Care | 46 |
| Gynaecological Oncology | 57 |
| Gynaecological Problems | 209 |
| Management of Delivery | 30 |
| Management of Labour | 45 |
| Maternal Medicine | 171 |
| Postoperative Care | 23 |
| Postpartum Problems | 32 |
| Sexual & Reproductive Health | 33 |
| Subfertility | 36 |
| Teaching & Research | 43 |

Supplementary Figure 2: Distribution of Number of Assessed MRCOG Part Two Questions by Knowledge Area.
